# PathoSPOT genomic epidemiology reveals under-the-radar nosocomial outbreaks

**DOI:** 10.1101/2020.05.11.20098103

**Authors:** Ana Berbel Caban, Theodore R. Pak, Ajay Obla, Amy C. Dupper, Kieran I. Chacko, Lindsey Fox, Alexandra Mills, Brianne Ciferri, Irina Oussenko, Colleen Beckford, Marilyn Chung, Robert Sebra, Melissa Smith, Sarah Conolly, Gopi Patel, Andrew Kasarskis, Mitchell J. Sullivan, Deena R. Altman, Harm van Bakel

## Abstract

Whole genome sequencing (WGS) is increasingly used to map the spread of bacterial and viral pathogens in nosocomial settings. A limiting factor for more widespread adoption of WGS for hospital infection prevention practices is the availability of standardized tools for genomic epidemiology. Here we present the Pathogen Sequencing Phylogenomic Outbreak Toolkit (PathoSPOT), which automates integration of genomic and medical record data for rapid detection and tracing of nosocomial outbreaks. To demonstrate its capabilities we applied PathoSPOT to complete genome surveillance data of 197 MRSA bacteremia cases from two hospitals during a two-year period. PathoSPOT identified 8 clonal clusters encompassing 33 patients (16.8% of cases); none of which had been recognized by standard practices. The largest cluster corresponded to a prolonged outbreak of a hospital-associated MRSA clone among 16 adults, spanning 9 wards over a period of 21 months. Analysis of precise timeline and location data with our toolkit suggested that an initial exposure event in a single ward led to infection and long-term colonization of multiple patients, followed by transmissions to other patients during recurrent hospitalizations. Overall, we demonstrate that PathoSPOT genomic surveillance enables detection of complex transmission chains that are not readily apparent from epidemiological data and that contribute significantly to morbidity and mortality, enabling more effective intervention strategies.

## Introduction

The utility of whole genome sequencing to track transmissions and outbreak events is well-established, in particular for highly clonal pathogens such as *S. aureus*, where classical molecular typing methods such as multi-locus sequence typing (MLST) and pulsed-field gel electrophoresis (PFGE) do not provide enough resolving power [1-4]. Despite the increased use of WGS, bottlenecks remain that complicate its use in managing nosocomial outbreaks. Comparative genome analyses often require specialized knowledge and/or selection of appropriate reference sequences. Analysis and visualization frameworks are available to aid genome analysis in global or regional outbreaks [5-7], but these are less suited for nosocomial settings where genomic data need to be integrated with detailed patient histories for contact tracing. This can be time-consuming, especially when relying on manual chart review. Integration with electronic medical record (EMR) systems can aid this process, but tools that combine patient and genomic information in a comprehensive manner are not readily available.

To facilitate detection and mapping of transmission chains in nosocomial settings we developed the open-source <u>Patho</u>gen <u>S</u>equencing <u>P</u>hylogenomic <u>O</u>utbreak <u>T</u>oolkit (PathoSPOT), which combines automated comparisons of complete or draft genomes with interactive visualization of clonal clusters. Further integration of epidemiological data enables high resolution analysis of outbreak phylogenies and contact tracing. We used our toolkit as part of a complete genome surveillance program of methicillin resistant *Staphylococcus aureus* (MRSA); a common cause of healthcare-associated infections in the USA posing a fatal threat to patients. PathoSPOT comparisons of MRSA genomes from 197 bacteremic patients identified multiple transmission events and a hospital-wide outbreak encompassing 16 patients that had not been detected by conventional infection prevention strategies. In-depth analysis with PathoSPOT allowed us to reconstruct the outbreak timeline and identify common links among these individuals. Our findings demonstrate the utility of PathoSPOT for precision surveillance in healthcare systems and highlight the role of colonization in long-term nosocomial outbreaks.

## Results

### PathoSPOT surveillance identifies frequent under the radar MRSA transmissions

We developed PathoSPOT to automate comparisons of large numbers of complete or draft microbial genomes, and to rapidly identify closely related isolates indicative of transmission events and map their epidemiological timelines (**Fig. 1A**). PathoSPOT combines existing tools for whole genome alignment with custom analysis and visualization code developed in Ruby, Python, and Javascript (https://pathospot.org). To demonstrate its utility, we applied PathoSPOT to MRSA isolates obtained from all bacteremia cases at two hospitals (A and B) during a 2-year period. In total we sequenced 224 genomes for 221 isolates from 197 patients using PacBio long-read technology and obtained 184 finished-quality and 40 draft genome sequences (**Table S1**). In most cases we only sequenced the primary blood culture, but additional isolates were analyzed for the same patient in cases of prolonged or recurrent infections. We first used the PathoSPOT "compare” pipeline to cluster genomes based on Mash distance [8]. This step groups related genomes prior to multi-genome alignment and avoids the need for manual selection of a reference genome. The Mash distance threshold for MRSA was determined empirically to yield clusters of genomes consistent with known clonal complex assignments based on MLST data, and to maximize core genome alignments (**Fig. 1B**). Pairwise distances between genomes were then calculated as the number of single nucleotide variants (SNVs) between core genome alignments in each cluster for further analysis.

**Figure 1.**
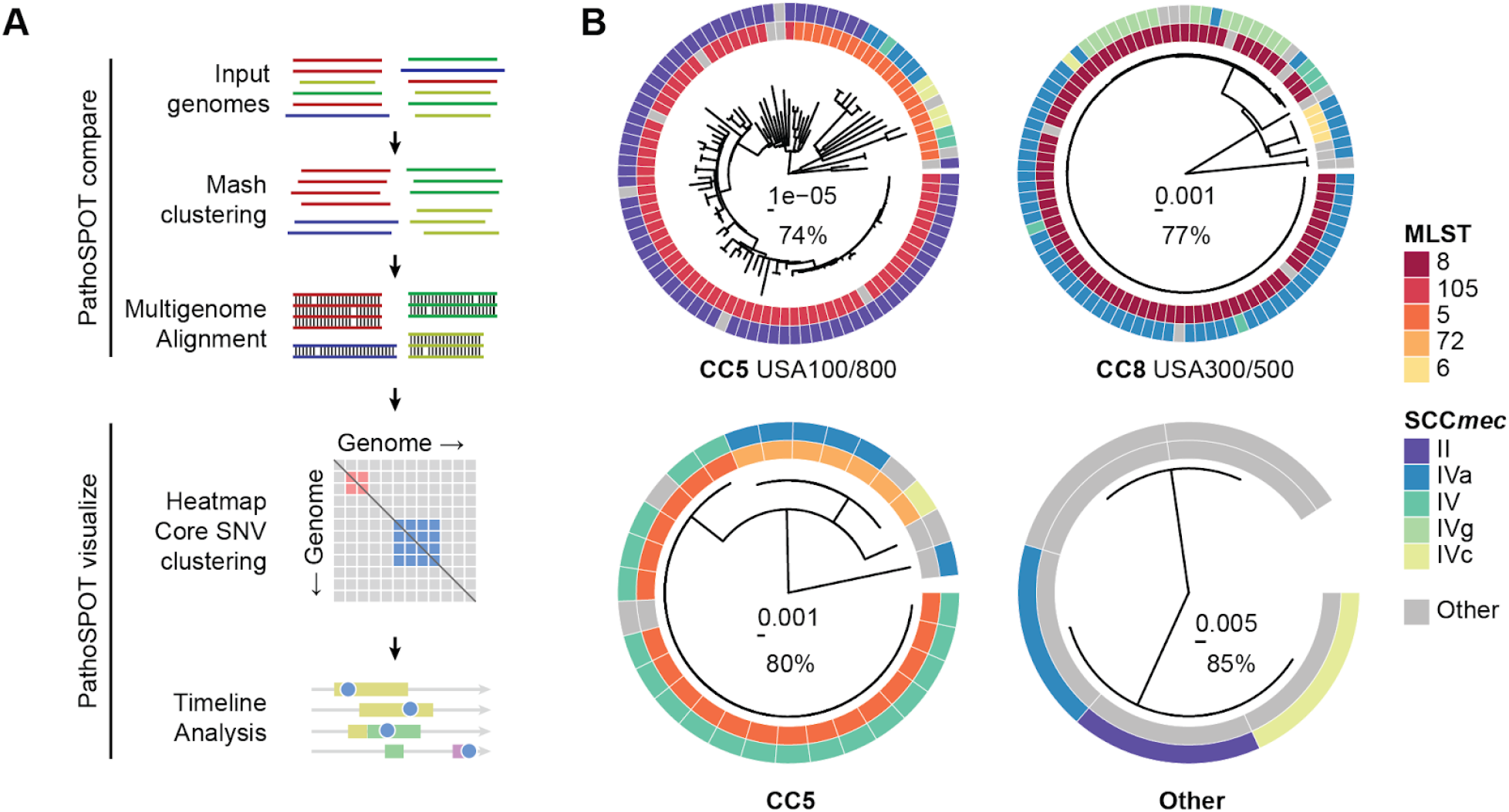
PathoSPOT comparative genome analysis of 221 MRSA isolates. **A)** Overview of the PathoSPOT whole-genome comparison framework. **B)** Maximum-likelihood phylogenetic trees produced from core genome SNVs identified from Parsnp whole-genome alignments of 4 clusters identified at a Mash score threshold of 0.02. Trees are annotated with MLST and *SCCmec* information (key shown on right) and clonal complexes (bottom). Scale bars indicating the number of substitutions per site in the phylogeny and the percentage of core genome coverage among all sequences is shown at the center of each tree.

To identify transmission events we used the PathoSPOT “heatmap” tool (**Fig. 2**). We set a threshold of ≤15 SNVs to identify potential transmissions, based on the extent of intra- and inter-patient variability we previously observed in complete genome analysis of an extended outbreak [4], and considering a core genome mutation rate of ~3 SNVs per Mb per year [2,9]. The distance threshold can be varied interactively in the heatmap tool to explore grouping at different levels of relatedness, depending on the pathogen. At the selected threshold we identified 8 clonal clusters with a total of 33 patients, implicating 16.8% of surveilled patients in transmission events (**Fig. 2C**). Most clusters consisted of patient pairs (5/8) but there were 3 with more than two patients. Patients within each cluster typically had overlapping hospital visits (75%) and stayed in the same wards at some point during these visits (63%), but in many cases MRSA bacteremia was only found after they transferred to different wards. This likely contributed to the fact that none of the clusters could be recognized epidemiologically. The most striking example of this was a cluster of 24 isolates from 16 patients that were collected over a period of 21 months from 9 different wards.

**Figure 2.**
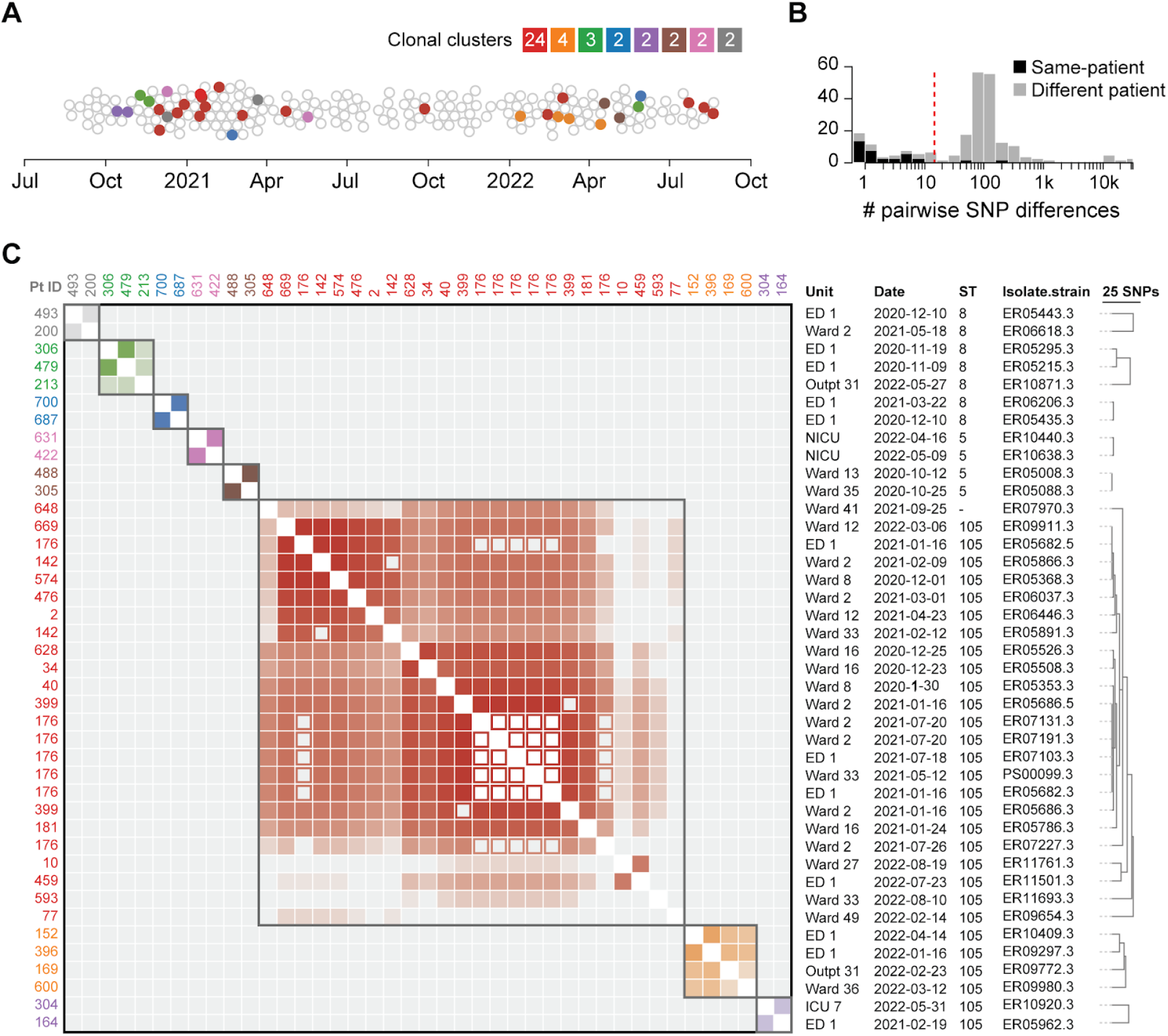
Identification of clonal clusters among 197 MRSA bacteremia cases. **A)** Beeswarm plot of MRSA cases with sequenced isolate genomes during the surveillance period. Cases with isolates separated by ≤15 core genome SNPs are grouped in clonal clusters, each highlighted with a distinct color. The number of isolates in each cluster is indicated in the color key. **B)** Histogram of pairwise core genome SNP distances for isolates obtained from the same patient (black bars) and isolates obtained from different patients (grey bars). The vertical red line indicates the 15-SNP threshold for clonality. **C)** Heatmap of pairwise core genome SNP distances between clustered isolates. Clusters are grouped along the diagonal and colored as in A, with decreasing shading reflecting an increased pairwise SNP distance. Closed squares and open squares are used for isolates from different patients or the same patient, respectively. All date information in this figure was recoded to protect health information.

### PathoSPOT timeline highlights the role of colonization in prolonged MRSA outbreaks

The presence of a clonal MRSA cluster among 16 bacteremia cases was consistent with a prolonged “under-the-radar” outbreak. The outbreak strain matched the hospital-associated USA100 lineage *(spa* type t002, MLST 105, staphylococcal cassette chromosome *mec* type II) and was resistant to fluoroquinolones, oxacillin, clindamycin, erythromycin and gentamicin. We next used the PathoSPOT “dendro-timeline” tool, which combines phylogenetic analysis of outbreak isolates with the admission/transfer/discharge (ADT) history for each patient, to analyze this outbreak in more detail (**Fig. 3**). The core genome dendrogram, derived from the multigenome alignment of the superset of isolates in the same Mash cluster, indicated the presence of distinct subclades within the outbreak (**Fig. 3A**). Shared variant profiles within each subclade were consistent with sub-transmissions within the larger outbreak (**Fig. 3A**). Isolates from patient 176 (p176), who tested positive for MRSA bacteremia numerous times within a span of 6 months, were represented in distinct clades, suggesting that the patient carried distinct variants of the outbreak strain at the same time. This was confirmed by sequencing two subclones from p176 isolate ER05682 (**Fig. 3A**, ▲ and ♦).

**Figure 3.**
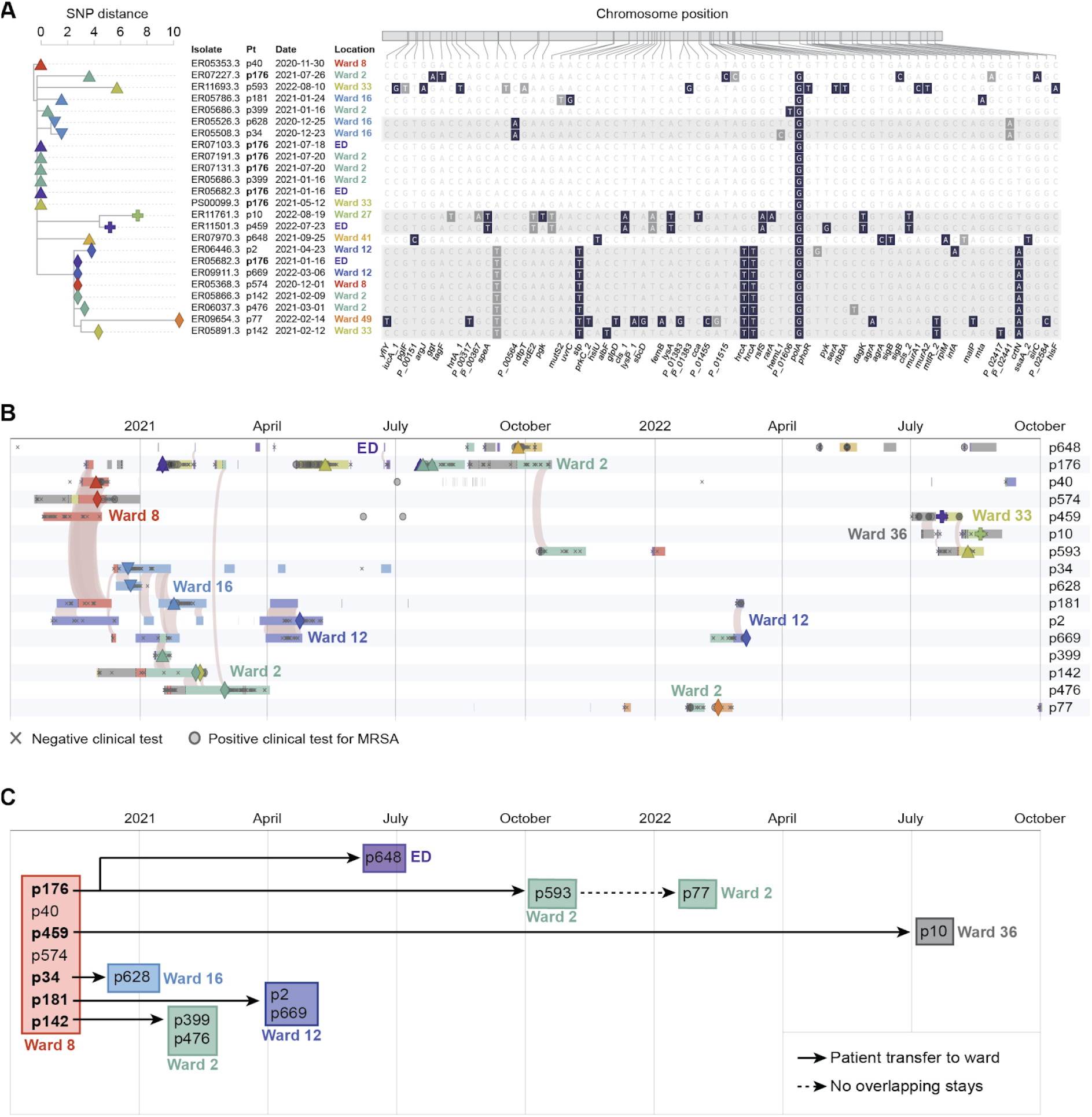
Epidemiologic timeline of long-term outbreak. **A)** Phylogenetic tree of core genome SNP differences (left) with corresponding locations of each variant relative to the first outbreak isolate that was obtained from p40 (right). Non-synonymous and synonymous variants are highlighted in black and grey, respectively. Distinct clades with three or more shared variants are shown as shaded areas. **B)** PathoSPOT timeline integrating genomic and epidemiological data. Rows correspond to patients, with admission periods in hospital wards shown as horizontal bars, colored as in A. Sequenced isolates are shown as different symbols matching those used in the phylogenetic tree in A. Shaded arcs signify ward-level patient overlap within 24 hours. Other positive or negative clinical test results are indicated by grey symbols, with a key shown below. The scale is shown at the top. **C)** Summary of key outbreak events in wards, derived from the epidemiological timeline. See main text for details. All date information in this figure was recoded to protect health information.

Based on the PathoSPOT timeline of events we reconstructed patient contacts based on location overlaps (**Fig. 3B**) and inferred the most likely outbreak scenario (**Fig. 3C**). The first two patients (p40 and p574) tested positive on ward 8. No other positives were found on this ward, but five other patients had overlapping stays (p176, p459, and p181) or were admitted to the same ward within four weeks of the first positive test (p34, p142). All but one patient tested positive for bacteremia within seven weeks of their stay in ward 8, following transfers or readmissions to other wards. Strikingly, p459, who was discharged from ward 8 four days after the initial positive case, did not present with bacteremia until 20 months later. In the intervening period this patient had no contact with our health system except for two dermatology office visits where positive wound cultures for MRSA were obtained. The degree of genetic drift of the p459 isolate (11 SNVs) and pattern of positive wound cultures prior to bacteremia are consistent with long-term colonization after initial exposure in ward 8, although we could not verify this scenario as the wound isolates were not available for sequencing.

Five additional patients tested positive in wards 16, 2, and 12 during the first six months of the outbreak (**Fig. 3B, 3C**). Each instance was preceded by the transfer of a patient that had previously stayed in ward 8, suggesting that direct or indirect transmissions from these cases propagated the outbreak. Notably, p34 was transferred from ward 8 to ward 16, into a room neighboring p628, who became bacteremic two days later. Both their isolates were grouped in the same subclade (**Fig. 3A**, ▼). Likewise, p142 was transferred from ward 8 to ward 2, where there was overlap with p399 and p476 before all three became bacteremic on this ward. Notably, p142 and p176 overlapped with p476 on two different days in the inpatient hemodialysis unit, providing an alternative acquisition route. Finally, after discharge from ward 8, p181 was readmitted to ward 12, where the patient overlapped stays with patients p2 and p669.

Four late transmission events were identified in months 7 to 21. Two of these events involved patient p176, who tested positive for the outbreak strain on multiple occasions during readmissions. P176 visited the emergency department (ED) on the same day as patient p648, and had an overlapping stay in ward 2 with p593 for at least 5 days, in the months prior to their positive tests. Following readmission after a 20-month hiatus, p459 likely transmitted to p10, based on evidence of an overlapping stay in ward 36 (**Fig. 3B**) and the high relatedness of their isolates (**Fig 3A**, +). Patient p77 was the only person that did not have overlapping stays with other outbreak cases. The patient had a total of two pediatric (ward 49) and one adult (ward 2) admission to the hospital within 21-months. Given that all other outbreak cases were adults, we consider ward 2 the most likely location of MRSA acquisition, where p77 shared healthcare workers with p593 who was admitted to the same unit 11 weeks before.

Altogether, PathoSPOT analysis suggested that initial exposure in Ward 8 resulted in colonization and subsequent clinical infection of 7 patients (44% of the prolonged outbreak cluster), followed by secondary transmissions after ward transfers and/or readmission of these initial cases. An alternative scenario of community transmissions was discounted after mapping of home zip codes, which indicated that 13 of 16 cases lived in geographically distinct neighborhoods. Spatiotemporal analyses of the seven smaller clusters (**Fig. S1**) showed that five included direct overlaps, of which three are plausible transmission events, and two such events occurred months before the clonal blood cultures were obtained.

### Hand hygiene compliance and vascular access implicated in under-the-radar outbreak

As the outbreak extended over multiple wards, we further investigated hand hygiene rates, shared HCW, patient movements, and clinical characteristics. Average hand hygiene compliance in affected wards ranged between 79-83%. Compliance in wards 8 and 16 decreased to 70% and 66% in the month prior to the first outbreak case, while ward 2 compliance was maintained at 79%. All outbreak patients shared at least one HCW involved in the care of other patients in the cluster. This is consistent with the high degree of overlapping stays in the same ward and suggests that direct and indirect transmissions may have played a role in propagating the outbreak. Although outbreak cases were moved frequently between units based on transfer records, they did not move more frequently than non-outbreak patients.

Chart review and univariate and multivariate analyses of the 16 outbreak cases compared to 34 patients infected with non-outbreak MLST 105 MRSA showed that outbreak cases were significantly associated with HO-MRSA (OR= 5.20 95% CI [1.04-26.01]; p=0.04) and intravenous chemotherapy prior to bacteremia (OR=11.24 95% CI [1.72-73.28]; p=0.01) (**Table 1**). Notable was the presence of active malignancy (57%; n=8) including leukemia (n=5), multiple myeloma (n=1), disseminated Kaposi sarcoma (n=1), and metastatic breast cancer (n=1). Consistent with these findings, the most common presumed source of bacteremia was vascular access (n=9; 56%), followed by skin source (n=4; 25%). The 90-day mortality incidence was 25% (n=4), of which 75% (n=3) was related to bacteremia with outbreak strain. There were no differences in outcomes between outbreak and non-outbreak patients.

**Table 1.** Outbreak patients vs. non-outbreak patients with MLST 105 isolates.

| Factor | Outbreak Patients<br>N = 16 (%) | Non-Outbreak Patients<br>N = 34 (%) | Univariate Analysis<br>OR (95% CI) | p value | Multivariate Analysis<br>OR (95% CI) | p value |
| --- | --- | --- | --- | --- | --- | --- |
| Male | 11 (69) | 21 (62) | 1.36 (0.39-4.82) | 0.63 |  |  |
| *Race/Ethnicity* |  |  |  |  |  |  |
| Non-Hispanic White | 5 (31) | 14 (41) | Reference |  |  |  |
| Non-Hispanic Black | 3 (19) | 9 (26) | 0.93 (0.18-4.90) | 0.94 |  |  |
| Hispanic/Latino/Asian | 5 (31) | 6 (18) | 2.33 (0.49-11.17) | 0.29 |  |  |
| Unknown | 3 (19) | 5 (15) | 1.68 (0.29-9.75) | 0.56 |  |  |
| *Age at Time of Infection* |  |  |  |  |  |  |
| 18-54 Years | 6 (38) | 7 (21) | Reference |  |  |  |
| 55-69 Years | 5 (31) | 9 (26) | 0.65 (0.14-3.04) | 0.58 |  |  |
| ≥ 70 Years | 5 (31) | 18 (53) | 0.32 (0.07-1.41) | 0.13 |  |  |
| History of IV Drug Use | 2 (13) | 2 (6) | 2.29 (0.29-17.90) | 0.43 |  |  |
| HIV | 1 (6) | 3 (9) | 0.69 (0.07-7.19) | 0.76 |  |  |
| *Admission Source* |  |  |  |  |  |  |
| Home | 12 (75) | 17 (50) | Reference |  |  |  |
| NH/Rehab/LTACH | 2 (13) | 11 (32) | 0.26 (0.05-1.38) | 0.11 |  |  |
| Other Hospital | 2 (13) | 6 (18) | 0.47 (0.08-2.75) | 0.40 |  |  |
| Prior Hospital Admission (90 Days) | 10 (63) | 23 (68) | 0.80 (0.23-2.76) | 0.72 |  |  |
| *NHSN Definitions* |  |  |  |  |  |  |
| CO-MRSA | 4 (25) | 24 (71) | Reference |  | Reference |  |
| HO-MRSA | 12 (75) | 10 (29) | **7.20 (1.87-27.79)** | **0.004** | **5.20 (1.04-26.01)** | **0.04** |
| <sup>A</sup> Presence of Invasive Device | 14 (88) | 27 (90) | 0.78 (0.12-5.21) | 0.80 |  |  |
| <sup>B</sup> Receiving Cancer Treatment | 7 (44) | 2 (6) | **15.75 (1.75-141.39)** | **0.01** | **11.24 (1.72-73.28)** | **0.01** |
| *Charlson Comorbidity Index (CCI)* |  |  |  |  |  |  |
| 0-3 | 4 (25) | 9 (26) | Reference |  |  |  |
| ≥ 4 | 12 (75) | 25 (74) | 1.08 (0.28-4.23) | 0.91 |  |  |
| History of MRSA Colonization | 6 (38) | 16 (47) | 0.68 (0.20-2.28) | 0.53 |  |  |
| *Presumed Source of MRSA BSI* |  |  |  |  |  |  |
| <sup>C</sup> Skin Source | 4 (25) | 7 (21) | 1.29 (0.32-5.24) | 0.73 |  |  |
| Pneumonia | 1 (6) | 6 (18) | 0.31 (0.03-2.83) | 0.30 |  |  |
| <sup>*</sup>, <sup>†</sup> Vascular access | 9 (56) | 9 (26) | **3.57 (1.03-12.43)** | **0.05** |  |  |
| Other/Undetermined | 2 (13) | 12 (35) | 0.26 (0.05-1.35) | 0.11 |  |  |
| Persistent Bacteremia (≥ 5 Days) | 2 (13) | 9 (26) | 0.40 (0.08-2.10) | 0.28 |  |  |
| ICU Admission Prior to BSI | 4 (25) | 3 (9) | 3.44 (0.67-17.73) | 0.14 |  |  |
| Intubated Prior to MRSA BSI | 4 (25) | 2 (6) | 5.33 (0.86-33.00) | 0.07 |  |  |

**Bold** = significant at ≤ 0.05
Abbreviations: BSI, bloodstream infection; HIV, human immunodeficiency virus; ICU, intensive care unit; IV, intravenous; NHSN, National Healthcare Safety Network.
<sup>A</sup> Includes devices such as pacemaker, any vascular access, orthopedic hardware, foley catheter, arteriovenous graft placement, percutaneous endoscopic gastronomy (PEG), ostomy, or any type of urinary collection at the time of first positive bloodstream infection.
<sup>B</sup> Includes patients actively receiving cancer treatment through a central venous catheter prior to bacteremia in the outpatient or inpatient setting.
<sup>C</sup> Skin source includes skin and soft tissue infections, thrombophlebitis due to peripheral IV catheters
<sup>\*</sup> Variable not included in the multivariate analysis in order to prevent collinearity between receiving cancer treatment and vascular access.
<sup>†</sup> Vascular access devices include: non-tunneled central venous catheter, tunneled catheter (hickman or permacath), implanted port, peripherally inserted central catheter (PICC line), as well as arteriovenous graft (AVG) and fistula (AVF).

## Discussion

We developed PathoSPOT as a key component of an ongoing genomics-based pathogen surveillance program to facilitate detection of outbreaks and transmissions. Application of the toolkit to surveillance data from 197 patients with MRSA bacteremia over a two-year period demonstrates the utility of our toolkit and shows that nosocomial transmissions are important sources of morbidity and mortality. We find that in the absence of genomic surveillance many nosocomial transmissions of MRSA go undetected by standard infection prevention practices, as they only result in clinically apparent infections weeks to months later.

An outbreak among 16 patients from distinct adult medicine wards spanned nearly the entire study period. Reconstruction of the epidemiological timeline with PathoSPOT suggests that this outbreak started with the exposure of 7 patients in a single ward. Subsequent transfers or readmissions of these patients to other wards were a key factor in propagating the outbreak across the hospital. Additional contributing factors may have included shared HCW and reduced hand hygiene rates surrounding key outbreak events. Frequent room changes within and between wards may also have resulted in contaminated environmental surfaces, which has been shown to play a role in the nosocomial transmissions [10,11]. In the absence of routine patient, HCW, and environmental screening it was not possible to determine the nature of the initial exposure event. The extended time between apparent exposure and subsequent bacteremia for most cases likely played a role in obscuring the outbreak from epidemiological detection.

Our study provides additional support for a role of colonization in the persistence and delayed progression of under-the-radar outbreaks [12,13]. Skin colonization in particular may have contributed to later infection, as vascular access was significantly associated as the presumed source of bacteremia among outbreak patients. The number of outbreak patients with hematological malignancies and bone marrow suppression was also notable in this respect. These patients are at an increased risk of bacteremia, as central venous catheters remain an essential tool for their treatment, frequently leading to catheter-related infections [14].

The detection of nosocomial transmissions and outbreaks is critical for healthcare organizations, and our findings have important ramifications for increasing the effectiveness of infection prevention strategies. Precise, genomics-based, pathogen surveillance programs supported by rapid analysis frameworks such as PathoSPOT are essential to detect events that are not readily ascertained by conventional epidemiological approaches. Widespread adoption of such programs depend on the availability of accessible tools that can be used by infection prevention staff without the need for extensive training. The effectiveness of such programs can be further increased when implemented across regional health systems, long-term acute care hospitals, and skilled nursing facilities, to track dissemination of strains and identify sources and at-risk patients based on contact networks. When combined with timely intervention, these efforts may be of critical importance to help reduce endemic rates of nosocomial infections.

## Methods

### Ethics statement

This study was reviewed and approved by the Institutional Review Board of the Icahn School of Medicine at Mount Sinai.

### Bacterial culturing, DNA extraction and sequencing

Isolates were cultured and identified as part of standard clinical testing procedures in the Mount Sinai Hospital Clinical Microbiology Laboratory (CML), and stored in tryptic soy broth (TSB) with 15% glycerol at −80°C. Selected isolates were subcultured on tryptic soy agar (TSA) plates with 5% sheep blood (blood agar) (ThermoFisher Scientific) under nonselective conditions. The Qiagen DNeasy Blood & Tissue Kit (Qiagen) was used for DNA extraction, as previously described [4]. Following quality control, DNA and library preparation, long-read sequencing was performed on the Pacific Biosciences RS-II platform to a depth of >200x.

### Genome assembly

PacBio SMRT sequencing data were assembled using a custom genome assembly and finishing pipeline (https://github.com/powerpak/pathogendb-pipeline), as previously described [4].

### Comparative genome analysis using PathSPOT-compare

We developed the PathoSPOT-compare pipeline to perform comparative phylogenomic analysis of finished, annotated genome assemblies for the specific purpose of outbreak detection. The pipeline is implemented as a Rakefile (a Makefile for the Ruby language) that calculates dependencies and executes all necessary subtasks to reach desired outputs. PathoSPOT-compare takes FASTA-formatted genome assemblies as input, along with a relational database (SQLite or MySQL) containing metadata for each assembly (including collection time, location, collection method, organism, and patient ID), as well as metadata on patient ADT history (for spatiotemporal analysis).

Genetic distances for outbreak detection are ultimately calculated by counting SNP differences within core genome alignments; however, there is a trade-off between aligning increasingly diverse assemblies and a diminishing core genome size (as more subsequences will fail to align across all assemblies). Therefore, we implemented a hybrid approach, wherein pairwise distances between all assemblies are first estimated using Mash [8], which uses a k-mer based hashing approach that approximates average nucleotide identity (ANI). Mash distances are used to perform greedy single-linkage hierarchical clustering, with clusters capped at a pre-specified diameter and size. The default parameters, which are also the parameters used for this study, are a maximum Mash cluster diameter of 0.02 (approximating 98% ANI among all included genomes) and at most 100 genomes per cluster.

Rapid core genome alignments are then created for each cluster using parsnp [15], which is tailored for intraspecific genome analysis and is therefore well-suited for outbreak analysis. Outputted variant call files (VCF) for each cluster are converted to NumPy arrays (NPZ files) for fast loading and subsetting of variant data by PathoSPOT-visualize, the downstream visualization web application that can display called variants alongside phylogenies. The primary output for PathoSPOT-visualize is a JSON file containing a matrix of pairwise SNP distances for all genomes (with inter-cluster distances left unspecified) and a maximum-likelihood phylogeny for each cluster. Additional optional pipeline tasks export patient location data (as TSV files) and epidemiologic data on positive and negative culture results (as JSON files), both of which are automatically utilized and layered onto the comparative genomic analyses within PathoSPOT-visualize when available.

### Interactive detection and visualization of outbreaks with PathoSPOT-visualize

To visualize the above analyses as depicted in **Fig. 2, 3A-B, S1, and S2**, we created the PathoSPOT-visualize interactive web application. The application uses PHP scripts and AJAX to serve data from the JSON, TSV, and NPZ output files generated by the PathoSPOT-compare pipeline, which are then dynamically mapped to interactive HTML5 and scalable vector graphics (SVG) elements using the D3.js (Data-Driven Documents) framework. All views are rendered in the browser, allowing the user to alter settings that trigger live animated transitions, and an intuitive sense of how changes propagate between the linked views of data.

There are three main user interfaces, the “heatmap” tool, the “network map” tool and the “dendro-timeline” tool. Users initially interact with the “heatmap” tool, which starts with selection of a dataset that can be prefiltered by specimen location, MLST, and time interval. The user can dynamically adjust the SNP threshold that specifies the genetic distance deemed suspicious for transmission; this process is aided by a histogram depicting distributions of SNP distances among same-patient isolates (generally expected to be related) and different-patient isolates (which are not, assuming a low level of transmission). This threshold is used to perform single-linkage hierarchical clustering of genomes on the client-side, with the clusters assigned random colors and depicted on a beeswarm timeline plot and a large heatmap of pairwise distances among all selected genomes. This large heatmap can be swapped for the “network map” view (**Fig. S2**), which plots genomes by their collection location in a geospatial layout, overlaid with density plots of overall epidemiological incidence and force-directed network links depicting genetic relationships.

Epidemiological links within the clusters can be further explored in the “dendro-timeline” tool, which combines a traditional phylogenetic dendrogram with a SNP matrix, a mapping of SNP locations onto a reference assembly, and a pannable-zoomable timeline of patient locations over time, with spatiotemporal overlaps highlighted as bright arcs. The phylogeny for the “dendro-timeline” tool is extracted from the larger maximum-likelihood trees built by parsnp, based on the SNP threshold and clustering parameters that the user selected in the “heatmap” tool.

### Case review

We performed a retrospective clinical chart review on all adults (age ≥18) subjects identified with MRSA bacteremia. Analyses were performed in SAS (v9.4)[16]. Variables were analyzed initially in a univariate logistic regression model. Variables *p*≤0.2 were then placed into a stepwise multivariate logistic regression model [17]. An additional in-depth chart review was performed for subjects identified as being involved in transmission events. These details included location (ward, room, bed), all ADT information, procedures, and providers. Whole genome sequencing was performed on 1st patient blood isolate positive for MRSA as part of an ongoing genomic surveillance program as described in [4]. Hand hygiene is monitored by the Infection Prevention and Control department by the use of anonymous observers using the Joint Commision’s Targeted Solutions Tool (TST) [18], which was implemented in November 2014. This tool allows the staff member to document reasons for non-compliance and target areas of interventions. Hand hygiene observations are collected anonymously at entry and exit by trained staff members in each hospital ward. Hand hygiene rates were compared using a one-sample t-test.

### Data availability

Genome assemblies have been deposited in Genbank (see Table S1 for accession numbers). All study data is also available at https://pathospot.org.

### Software

The PathoSPOT-compare and PathoSPOT-visualize packages developed for this study are both open source and can be obtained from:

- https://github.com/powerpak/pathospot-compare
- https://github.com/powerpak/pathospot-visualize

A live demo of all visualizations created for this study, along with documentation on setting up and using the software with example data from this study, can be found at https://pathospot.org.

## Data Availability

All study data is available at https://www.pathospot.org.

https://www.pathospot.org.

## Acknowledgements

This research was supported in part by R01 AI119145 (H.v.B.), the Icahn Institute for Genomics and Data Science (A.K.), the CTSA/NCATS KL2 Program (KL2TR001435, Icahn School of Medicine at Mount Sinai; D.R.A), and the New York State Department of Health Empire Clinical Research Investigator Program (Awarded to Judith A. Aberg, Icahn School of Medicine at Mount Sinai; D.R.A.) and F30 AI122673 (T.R.P.). The Research reported in this paper was supported by the Office of Research Infrastructure of the National Institutes of Health (NIH) under award numbers S10OD018522 and S10OD026880 as well as institutional funds. The funders had no role in study design, data collection and interpretation, or the decision to submit the work for publication.

## Supplemental Figures

**Figure S1.**
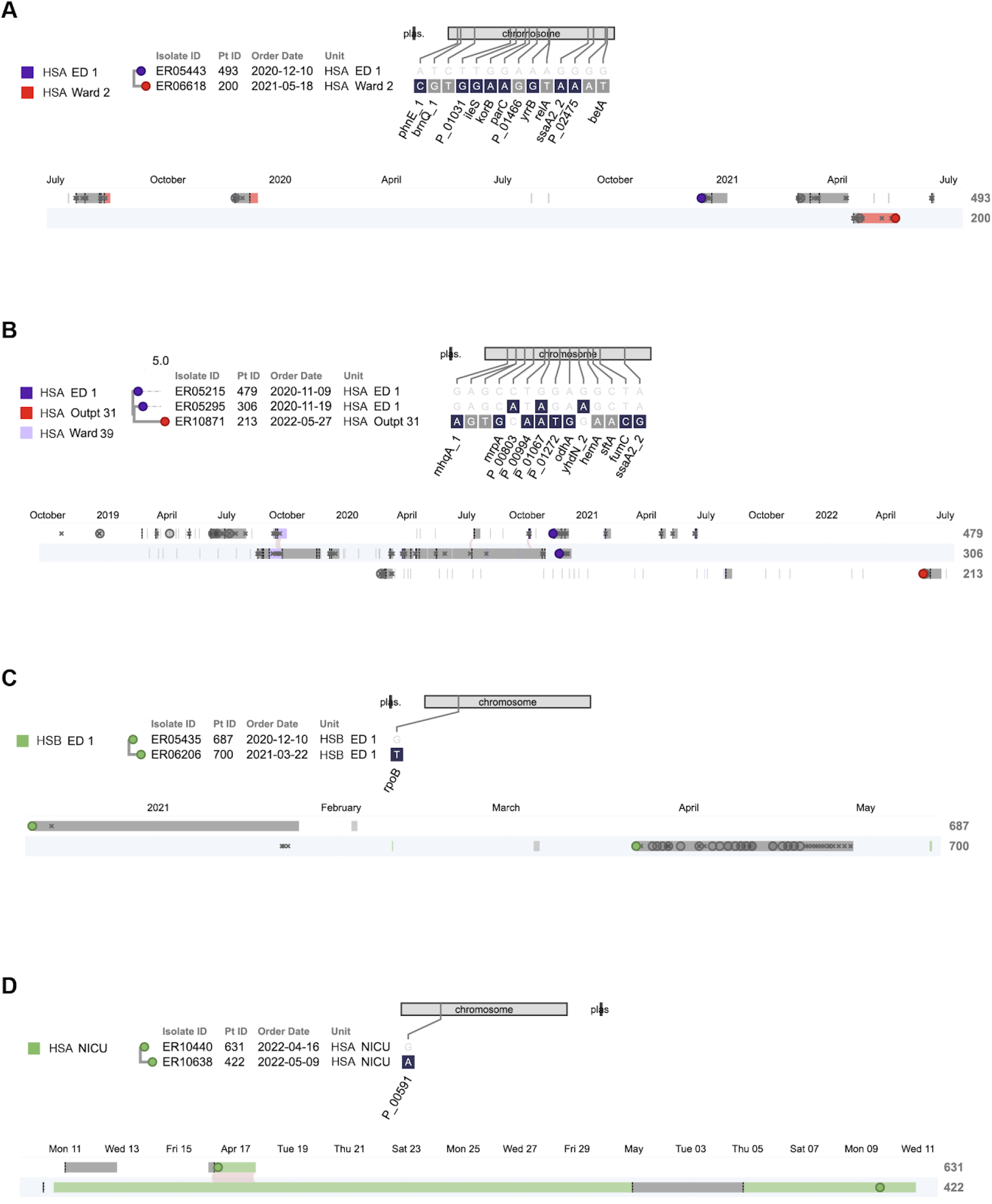
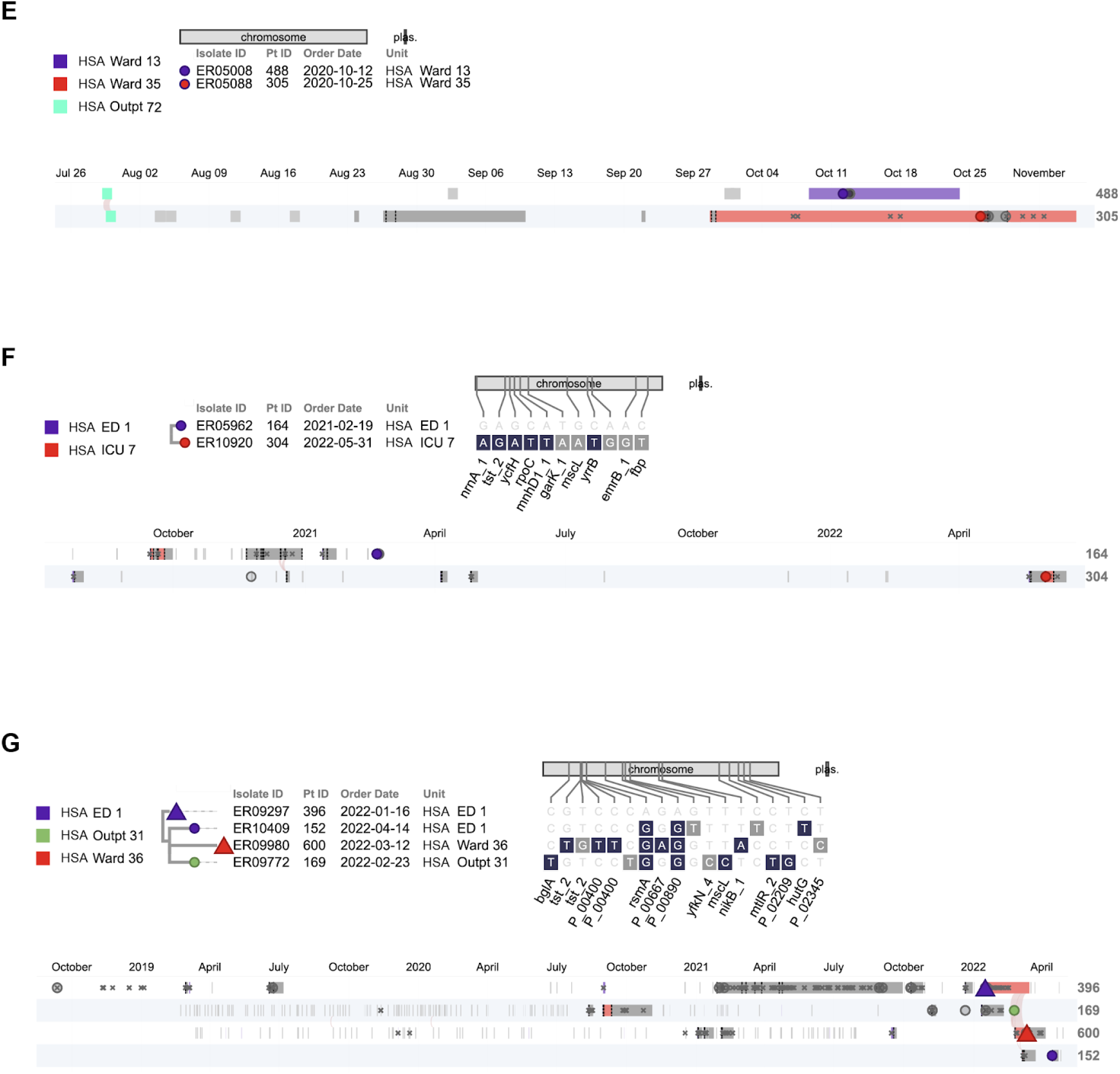
Other clonal clusters identified by PathoSPOT analysis. Phylogenetic trees of core genomes and epidemiologic timelines created by the “dendro-timeline” visualization in PathSPOT for the seven smaller clonal clusters identified in our study **(A-G)**. Layout and drawing conventions are as in Figures 3A and B. All date information in this figure was recoded to protect health information..

**Figure S2.**
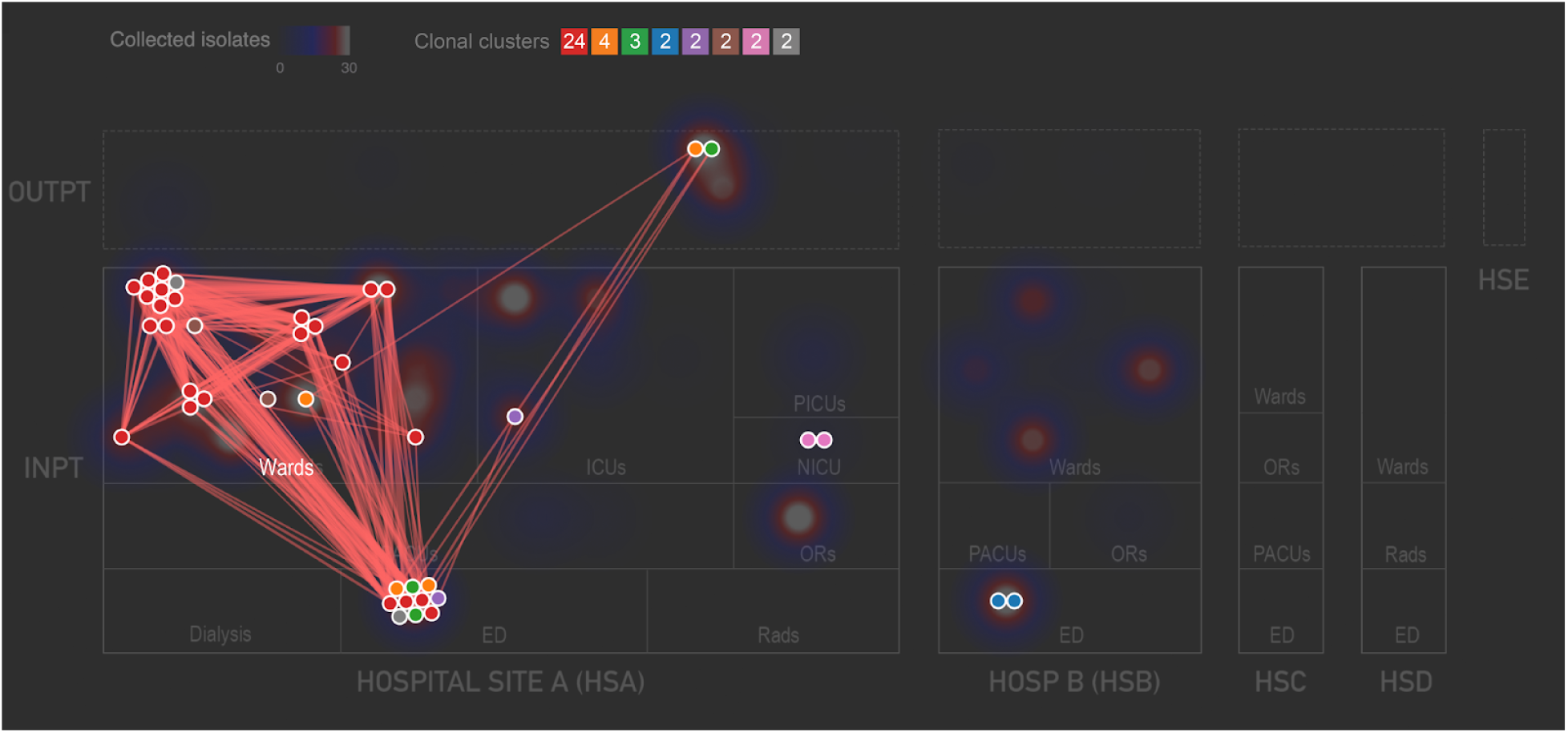
Network layout view of clonal clusters. The PathoSPOT network layout shows spatial relationships among patients in each of the 8 clusters, using the threshold of ≤15 SNVs across the entire 24 months of the dataset, and only the first related isolate from each patient. Nodes are colored by clonal cluster, as indicated in the legend at top center, which also lists the numbers of nodes in each cluster. Nodes are laid out spatially by hospital and ward where the patient was sampled, with ward types grouped together (labeled boxes), and nodes from the same ward placed adjacent to one another (the position of each anonymized ward within the box is arbitrary). Genomic links underneath the threshold are depicted as red lines, with many clusters spread across different wards and ward types. Underneath the network map, the total number of positive cultures collected from each location is depicted as a shaded density plot, with the color scale depicted at top left; this highlights heavily sampled locations.

